# Estimating productivity losses per HIV infection due to premature HIV mortality in the United States

**DOI:** 10.1101/2025.06.02.25328805

**Authors:** Md Hafizul Islam, Harrell Chesson, Ruiguang Song, Angela B. Hutchinson, Ram K. Shrestha, Alex Viguerie, Paul G. Farnham

## Abstract

**Background:** Updated estimates of the productivity losses per HIV infection due to premature HIV mortality are needed to help quantify the economic burden of HIV and inform cost-effectiveness analyses.

**Methods:** We used the human capital approach to estimate the productivity loss due to HIV mortality per HIV infection in the United States. We incorporated published data on age-specific annual productivity, life expectancy at HIV diagnosis, life years lost from premature death among persons with HIV (PWH), the number of years from HIV infection to diagnosis, and the percentage of deaths in PWH attributable to HIV. For the base case, we used 2018 life expectancy data for all PWH in the United States. We also examined scenarios using life expectancy in 2010 and life expectancy for cohorts on antiretroviral therapy (ART). We conducted sensitivity analyses to understand the impact of key input parameters.

**Results:** We estimated the base case overall average productivity loss due to HIV mortality per HIV infection at $65,300 in 2022 US dollars. The base case results showed a 45% decrease in the estimated productivity loss compared to the results when applying life expectancy data from 2010. Productivity loss was 83% lower for cohorts of PWH on ART compared to the base case scenario. Results were sensitive to assumptions about percentage of deaths attributable to HIV and heterogeneity in age at death.

**Conclusion:** This study provides valuable insights into the economic impact of HIV mortality, illustrating reductions in productivity losses over time due to advancements in treatments.

**Disclaimer:** The findings and conclusions in this report are those of the authors and do not necessarily represent the official position of the Centers for Disease Control and Prevention.

**Funding:** This work was done as regular official duties of the authors as employees of the Centers for Disease Control and Prevention.

**Conflicts of Interest:** All authors report no conflicts of interest.

**Previous Presentation:** Presented in part as a poster presentation at the ISPOR 2024 Conference, May 5-8, 2024, Atlanta, GA.

**Highlights:**

- Updated estimates of productivity losses per HIV infection due to premature HIV mortality can help assess the total economic burden of HIV in the United States.
- This study estimates productivity losses per HIV infection for overall, by sex, and by varying ages of HIV infection.
- Advancement in treatment has contributed to significant reduction in productivity losses due to premature HIV mortality in the United States over the past decade.

## Introduction

Productivity losses are defined as the value of loss in productivity that occurs due to illness, treatment, disability or premature mortality (i.e., death occurring before reaching the expected age of life expectancy).^1,2^ Productivity losses due to human immunodeficiency virus (HIV) occur when a person’s ability to work or to perform other valuable tasks is reduced or eliminated due to HIV-related illness, treatment, side effects of treatment, disability, and premature death. Productivity loss (indirect cost) and medical costs (direct cost) are two of the primary components of the economic burden of HIV infections in the United States as typically reported in cost of illness studies.^3–7^

Although HIV incidence has a declining trend over the past decade, in 2022, there were 31,800 new HIV infections in the United States.^8^ While recent studies estimated the average discounted lifetime direct medical cost per HIV infection ranging from $416,200 to $1,174,700 in 2022 US dollars (USD)^9–13^, estimates of the productivity loss per HIV infection in the United States are limited. In 2006, Hutchinson et al. estimated the productivity loss due to HIV-related mortality in the United States as $1,207,200 in 2022 USD.^14^. The authors estimated the HIV mortality-related productivity losses using a human capital approach, which incorporated published life expectancy estimates, wage income and household production estimates. However, this was published before the widespread use of HIV treatment, i.e., antiretroviral therapy (ART), does not reflect current HIV treatment practices, having been conducted before the introduction of immediate ART initiation.^15^ U.S. Food and Drug Administration (FDA) approved the first HIV medicine in 1987^16^, which was followed by the introduction of combined ART in 1996, recommendation of immediate ART initiation^15^, and more recently the introduction of long-acting injectable ART.^17^ According to the Centers for Disease Control and Prevention (CDC), by 2022, 65.1% of the PWH achieved undetectable viral load^18^, which has led to reduced HIV-related morbidity and mortality, and improved life expectancy among PWH.^15^

This study aims to update the productivity loss estimates, per HIV infection due to HIV-related premature mortality (hereafter referred to as HIV mortality) in the United States. We incorporated the most recent available life expectancy estimates among PWH, highlighting the impact of recent improvements in HIV treatment. We also compared the effects of universal ART use for which all PWH were treated with ART to current ART use in the US on productivity loss due to HIV mortality. These estimates can help quantify the economic burden of HIV in the United States and inform cost-effectiveness analyses of HIV prevention interventions.

## Methods and Input Parameters

### Methods

We estimated the productivity loss due to HIV mortality per HIV infection in the United States, discounted to the time of HIV infection. To estimate this productivity loss per HIV infection, we applied the human capital approach^19^, a common methodology for estimating productivity loss due to disease mortality/morbidity.^14,20–25^ In this approach, loss of life years are valued based on estimates of annual productivity, which include both market (paid) and non-market (unpaid) activities. Factors incorporated in the calculation of productivity loss per HIV infection include annual productivity (*P*), life expectancy at HIV diagnosis (*E*), number of life years lost due to premature death among PWH (*L*), years from HIV infection to diagnosis (*M*), and the percentage of deaths among PWH that are attributable to HIV (*Z*). Throughout this study, the term “life expectancy at HIV diagnosis” indicates the remaining life expectancy at the time of HIV diagnosis. In the subsequent sections, we present the detailed approach to incorporate these factors into the calculations of both undiscounted and discounted lifetime productivity loss per HIV infection.

#### Undiscounted productivity loss

The undiscounted lifetime productivity loss due to premature all-cause mortality in a person with HIV can be calculated as *P* × *L*, where *P* is the annual value of productivity per person in the United States and L is the number of years of life lost per PWH. In other words, someone with HIV who dies but would have otherwise lived additional L years can be thought of as losing *P* in annual productivity for those *L* years. Since the development of effective ART, life expectancy for PWH has been increasing, nearing that of people without HIV infection.^26^ In addition, not all premature deaths among PWH are attributable to HIV. As per the National HIV Surveillance System data, HIV-related death rates (i.e., mortality due to HIV) among people with diagnosed HIV decreased by 48.4% during 2010-2017.^27^ Therefore, to estimate the productivity loss due to HIV mortality per HIV infection, we adjust for the percentage of deaths in PWH that are attributable to HIV. If *Z* is the percentage of deaths in PWH that are attributable to HIV, then the average undiscounted productivity loss (*PL*) due to HIV mortality, per person with HIV can be approximated as *PL* = *P* × *L* × *Z*. However, as annual productivity varies by age, if a person with HIV dies at the age of *a*_*D*_, we can rewrite *PL* as 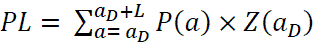, where *P*(*a*) is the productivity at age *a* and *Z*(*a*_*D*_) is the percentage of deaths at age *a*_*D*_ among PWH that are attributable to HIV.

#### Discounted productivity loss

The undiscounted productivity loss per HIV-related death is discounted to the time of HIV infection to estimate the discounted lifetime productivity loss at that point. We discounted the productivity loss in three steps: first to the time of death, then to the time of HIV diagnosis, and then to the time of HIV infection (Figure 1).

**Figure 1:**
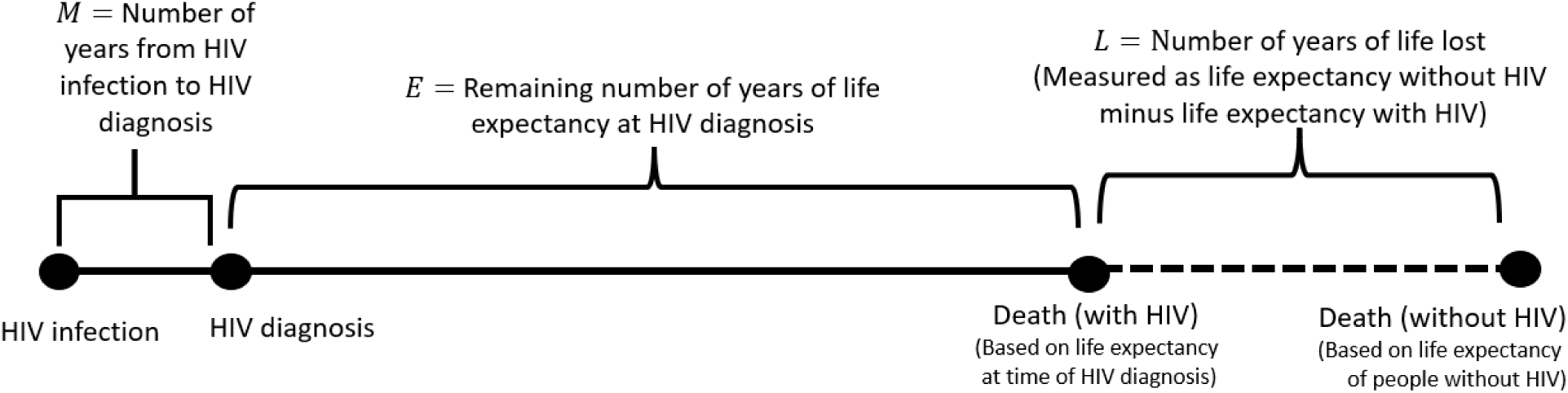
A schematic showing the calculation of the discounted productivity loss due to HIV mortality. Annual productivity losses were incurred over the years of life lost among people with HIV compared to people without HIV, shown by the dashed line. These productivity losses were discounted to the time of HIV infection at a rate of 3% annually, in three steps. First, the annual productivity losses shown by the dashed line were discounted to the time of death. Second, these productivity losses were further discounted to the time of HIV diagnosis. Third, these productivity losses were further discounted to the time of HIV infection.

The productivity loss due to HIV mortality discounted to the time of death (*PL_D_*) can be calculated as,

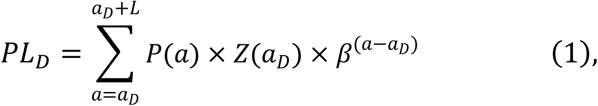

where β = 1/(1 + *r*) and *r* ≥ 0 is the annual discount rate.

In the second step, we discounted from the time of death to the time of HIV diagnosis. To do so, we discounted for additional *E*(*a*_*DX*_) years, multiplying *PL*_*D*_ in equation (1) by β^*E*(*aDX*)^, where *E*(*a*_*DX*_) is the number of years from HIV diagnosis to death with a HIV diagnosis at age *a*_*DX*_. Thus, the discounted productivity loss due to HIV mortality per HIV infection, discounted to the time of HIV diagnosis (*PL*_*DX*_) can be calculated as,

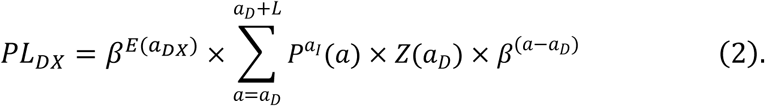

In the third step, we discounted from the time of HIV diagnosis to the time of HIV infection. To do this, we apply an additional discount for *M*(*a*_*DX*_) years, where *M*(*a*_*DX*_) is the number of years from HIV infection to diagnosis with an HIV diagnosis at age (*a*_*DX*_). To discount for *M*(*a*_*DX*_) years, we multiplied by β^*M*(*aDX*)^. Thus, the present value of the productivity loss due to HIV mortality per HIV infection, discounted to the time of infection (*PL*_*I*_) is expressed as,

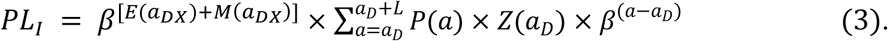

### Input Parameters

The input parameters that were used in the calculation of productivity loss estimates for the base case and other comparative scenarios are presented in Table 1 and are also discussed in detail below.

**Table 1:** Input parameters for base case and other comparative scenarios.

| Input parameters | All | Male | Female | Sources |
| --- | --- | --- | --- | --- |
| Median time (in years) from HIV infection to diagnosis ($M$) | | | | |
| Age: 13-34 | 3 |  |  | 39 |
| Age: 35-44 | 4 |  |  |  |
| Age: 45-54 | 5 |  |  |  |
| Age: 55+ | 6 |  |  |  |
| Percent of deaths attributable to HIV among PWH<sup>¥</sup> ($Z$) | 30% | | | 27 |
| Annual discount rate ($r$) | 3% | | | 45 |
| Remaining Life expectancy (in years) at HIV diagnosis ($E$) | | | | |
| PWH in 2018 (Base case) | 36.4 | 36.1 | 36.6 | 31 |
| PWH in 2010 | 31.1 | 31.7 | 30.4 | 32 |
| Cohort on ART<sup>§</sup>: initiated between 2015-2019 | 46.8 | 45.8 | 47.8 | 33 |
| Cohort on ART: initiated between 1996-2014 | 43.4 | 42.7 | 44.1 | 33 |
| Number of years of life lost among PWH ($L$) | | | | |
| PWH in 2018 (Base case) | 13.7 | 11.8 | 15.6 | 31 |
| PWH in 2010 | 18.9 | 16.1 | 21.6 | 32 |
| Clinical Trial Cohort on ART: ART initiated between 2015-2019 | 3.5 | 2.3 | 4.6 | 33,35 |
| Clinical Trial Cohort on ART: ART initiated<br>between 1996-2014 | 6.8 | 5.4 | 8.3 | 33,34 |

§ART = Antiretroviral therapy, ¶PWH = Persons with HIV

#### Annual productivity

We used age-specific annual mean productivity (market plus nonmarket) of US adults in 2016 US dollars reported by Grosse et al. 2019^28^, adjusted for inflation to 2022 USD using the “all items in US city average, all urban consumer, not seasonally adjusted, annual average” component of the US consumer price index, where the all items are categorized into 8 major groups, e.g., food and beverages, housing, apparel, transportation, medical care, recreation, education and communication, and other goods and services.^29^ Market productivity refers to paid labor, and nonmarket productivity refers to unpaid activities, such as providing childcare and eldercare, cleaning the home, and grocery shopping. Single age annual productivity data in 2022 USD for US adults aged 15-99 years for overall, male, and female are presented in supplementary material (Table S1). Productivity at each future age is determined by taking the 2022 productivity at that age and adjusting it to an annual growth rate of 1%, compounded over the age difference. Productivity differences for males and females primarily result from greater female absence in the workforce while bearing and raising children and slower earnings growth with increasing age for females.^28^

#### Life expectancy and number of life years lost

Median age of HIV diagnosis in the United States falls in the age group of 25-34 years.^30^ Hence, in our base case scenario, we used life expectancy after an HIV diagnosis at the age of 30 years, which is approximately the midpoint of the age range 25-34 years, and the corresponding years of life lost among people with diagnosed HIV, to estimate the productivity loss due to HIV mortality. Improved treatment and care have increased the life expectancy among PWH who are diagnosed early and who initiate and maintain treatment with antiretroviral therapy (ART). To understand the impact of improved life expectancy among PWH on the productivity loss due to HIV mortality, we obtained life expectancy data from different published studies. In addition to our base case estimate of the productivity loss due to HIV mortality per HIV infection using population-based 2018 life expectancy after HIV-diagnosis data, we also calculated the productivity loss due to HIV mortality per HIV infection using 2010 population-based life expectancy after HIV diagnosis data to illustrate trends in productivity loss due to HIV mortality over time due to improvements in HIV treatment. Similarly, we examined scenarios in which all people diagnosed with HIV initiated treatment with ART reflecting universal ART use compared to the population-based estimates that reflects current ART use in the US. We also used population-based 2018 life expectancy at varying ages of HIV diagnosis and the corresponding years of life lost compared to general population to estimate HIV mortality-related productivity loss at varying ages (for which life expectancy data were available) of HIV diagnosis.

##### US population-based HIV surveillance data

We used life expectancy after HIV diagnosis among adults and adolescents (13 years and older) in the United States from various time periods based on national HIV surveillance data. In the base case, we used population-based HIV surveillance data that reported the life expectancy of people diagnosed with HIV at age 30 years in 2018 as 36.4 years for all, 36.1 years for males, and 36.6 years for females (Table 1).^31^ Compared to the life expectancy of the general population at the age of 30 years, the corresponding years of life lost were 13.7 years for all, 11.8 years for males, and 15.6 years for females. To understand the trend in life expectancy, from a prior US population-based study restricted to persons with diagnosed HIV, we obtained life expectancy after HIV diagnosis at age 30 years in 2010 as 31.1 years for all, 31.7 years for males, and 31.4 years for females, and corresponding years of life lost in 2010 as 18.9 years, 16.1 years, and 21.6 years for all, males, and females, respectively (Table 1).^32^ Moreover, to estimate the productivity loss of from premature mortality for PWH at different ages, we used life expectancy of people diagnosed with HIV in 2018 at varying ages of HIV diagnosis and the corresponding life years lost from the population-based HIV surveillance data (Table S2).^31^

##### Clinical Trial Cohort on ART

To estimate the productivity loss for the scenario of all PWH receiving ART reflecting universal ART use, we used life expectancy data for PWH who remained on ART continuously for at least 1 year, as derived from clinical trial cohort studies conducted in Europe and North America.^33^ To be consistent with the base case scenario and also to understand the trend in life expectancy among PWH on long-term ART, we obtained life expectancy at age 30 years for PWH on ART who started ART from 1996–2014 and 2015–2019 by taking an average of life expectancy data reported for PWH at age 20 years and 40 years. The years of life lost due to any mortality for people diagnosed with HIV at age 30 years and on ART compared to the general population were calculated using life expectancy data for the US general population in 2014 and 2019 from the National Vital Statistics Report.^34,35^ Life expectancy after HIV diagnosis at age 30 years, both overall and by sex, along with the corresponding years of life lost for PWH receiving ART initiated between 1996–2014 and 2015–2019, are presented in Table 1.

#### Percentage of deaths in persons with HIV attributable to HIV

HIV attributable deaths among PWH have decreased since the introduction of ART. In our base case, we applied that 30% of deaths in PWH result from a known cause related to HIV, as reported in the literature for deaths among PWH at age 55 and higher based on data from National HIV Surveillance System.^27^ A 13-cohort study of patients on ART in Europe and North America from 1996 to 2006 estimated that 50% of deaths in PWH were due to HIV.^36^ We used this as an upper bound for the percentage of deaths among PWH that are attributable to HIV (*Z*). Based on the available data in 2021, approximately 25% of deaths among PWH in the United States were directly attributable to HIV.^37,38^ We used this as a lower bound for the percentage of deaths in PWH attributable to HIV.

#### Number of years from HIV infection to diagnosis

We applied median time from HIV infection to diagnosis (*M*) of 3 years for persons diagnosed with HIV at age 13-34 years, 4 years for age 35-44, 5 years for age 45-54, and 6 years for age 55 and over, based on estimates obtained from CDC’s National HIV Surveillance System data of patients diagnosed with HIV in 2018 in the United States.^39,40^

#### Sensitivity analyses

We conducted sensitivity analyses to examine how the estimates of the discounted productivity loss due to HIV mortality per HIV infection changed when we varied the percentage of deaths in people with HIV attributable to HIV (*Z*). We varied the parameters as per the upper and lower bounds presented in the input parameter section.

In our base case, for each person diagnosed with HIV at age 30 years, we applied the age at death as 66.4 years compared to 80.1 years for those without HIV. However, this simplification ignores the fact that some people diagnosed with HIV at age 30 years will die within a few years of diagnosis and some will live beyond age 66.4 years. To illustrate the impact of accounting for heterogeneity in age at death, we divided those diagnosed with HIV at age 30 years into 2 groups, one with shorter life expectancy after diagnosis (5 years) and the other with longer life expectancy after diagnosis (more than 5 years) and calculated the weighted average productivity losses of the two groups. We assumed 4.9% of people would be in Group 1, based on 4.9% 5-year mortality among people entering care for HIV in 2011-2017 as reported by Edwards et al. 2022.^41^ The age at death for those in Group 2 was calculated so that the weighted average age of death across Group 1 and Group 2 would be 66.4 years, the same as the average age of death in the base case when we assumed no heterogeneity in age at death.

## Results

We estimated an overall average productivity loss due to HIV mortality, discounted to the time of infection, after an HIV diagnosis at the age of 30 years as $65,300 (2022 USD) per HIV infection (Table 2). The average productivity losses due to HIV mortality per HIV infection for males and females were $67,400 and $63,300, respectively. When using life expectancy data from a 2010 US population-based study (instead of 2018 life expectancy estimates as in the base case), we estimated average lifetime productivity loss due to HIV mortality, discounted to time of infection, after an HIV diagnosis at age 30 years as $119,700, $119,000, and $118,900 for overall, males, and females, respectively.

**Table 2:** Productivity loss due to HIV mortality per HIV infection, discounted to the time of infection, among person with HIV (PWH) in the United States in the base case and other comparative scenarios (in 2022 US dollars).

| Scenario | Lifetime productivity loss due to HIV mortality<br>per HIV infection ($LPL_I$)* | | |
| --- | --- | --- | --- |
|  | All | Male | Female |
| PWH in 2018 (Base case)<sup>¥</sup> | \$65,300 | \$67,400 | \$63,300 |
| PWH in 2010 | \$119,700 | \$119,000 | \$118,900 |
| Clinical Trial Cohort on ART<sup>§</sup><br>(ART initiated between 2015-2019) | \$10,900 | \$8,500 | \$11,800 |
| Clinical Trial Cohort on ART<br>(ART initiated between 1996-2014) | \$24,200 | \$22,700 | \$24,900 |

\*Productivity loss estimates rounded to nearest \$100, <sup>¥</sup>To see the base case results stratified by market and nonmarket
productivity, see Supplemental Table S3. <sup>§</sup>ART = Antiretroviral therapy

In the base case, with 3 years of median time before HIV diagnosis and an overall life expectancy of 36.4 years among persons with HIV, a person diagnosed with HIV at age 30 (infected with HIV at age 27) is expected to continue being productive from the initial time of infection before diagnosis (from age 27-29) until death (approximately at age 66). Productivity loss attributable to HIV mortality is incurred for the number of years lost due to premature death due to HIV, from year 66 to year 80 of expected normal life (Figure 2). Productivity loss due to HIV mortality per HIV infection diagnosed at age 30 years for the cohort on ART (at least one year) was estimated to be $10,900 for ART initiated between 2015-2019 and $24,200 for ART initiated between 1996-2014.

**Figure 2:**
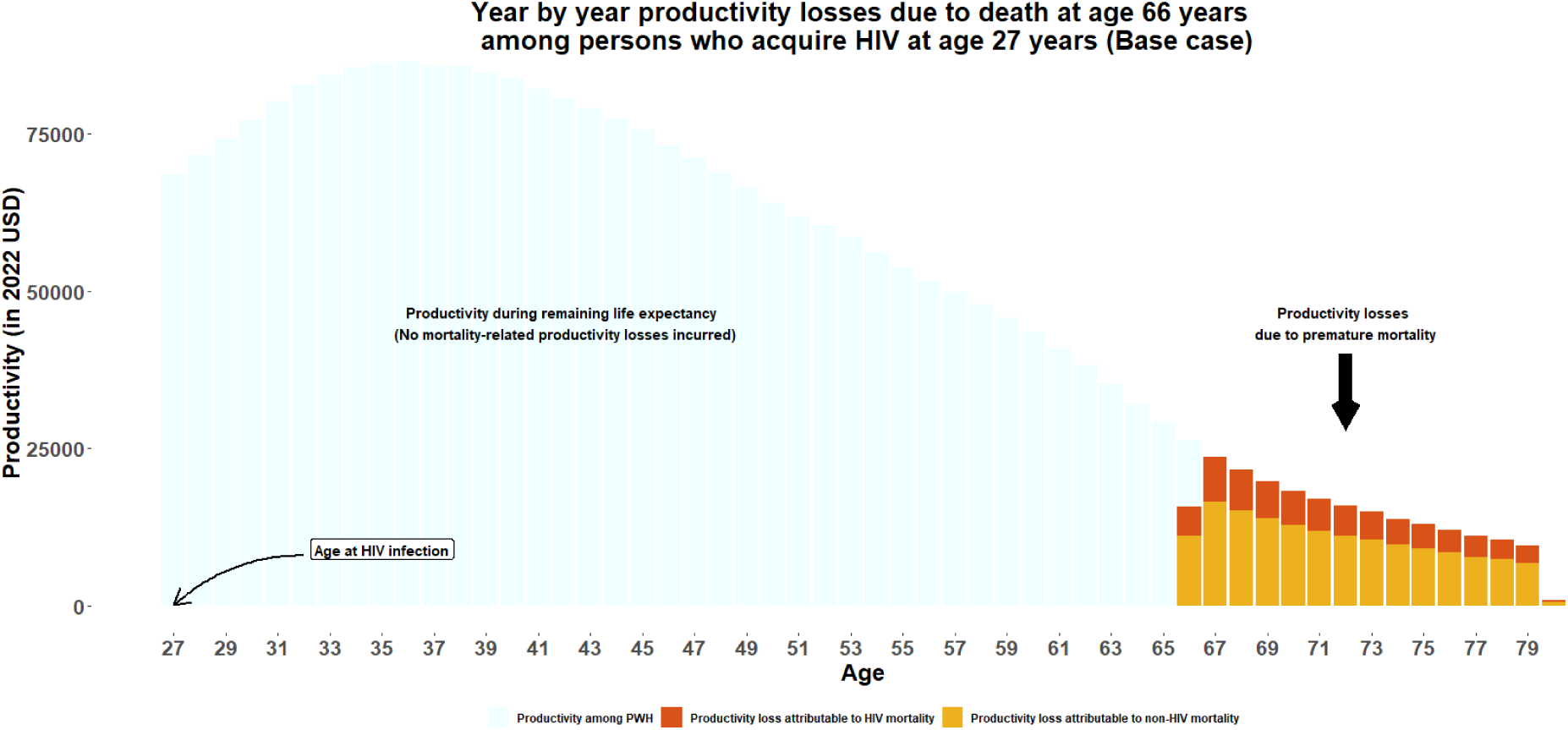
A year by year representation of the discounted productivity among people with HIV (PWH) and productivity loss due to HIV mortality discounted to the time of HIV infection for the base case scenario. A person acquiring HIV at age 27 years would be diagnosed with HIV at about age 30 years and would live an additional 36.4 years, 13.7 years fewer than the person would have lived without HIV. Thus, the person with HIV is assumed to be productive every year from the time of acquiring HIV until death around age 66 years. Productivity losses are incurred for the 13.7 additional years (∼age 67 years to ∼age 80 years) that the person would have lived, based on life expectancy among people without HIV. Because not all deaths in people with HIV are caused by HIV, only a portion of the productivity losses due to premature mortality in people with HIV are attributable to HIV (30% in the base case).

The estimated productivity loss due to HIV mortality per HIV infection, discounted to the time of infection, varied significantly by the age of HIV infection (Figure 3). Overall, an HIV infection at age 22, 27, and 32 years, correspondingly diagnosed at age 25, 30, and 35 years, was estimated to have the highest productivity loss ($65,300) due to HIV mortality (Table S3). The lowest estimated productivity loss ($4,300) due to HIV mortality, discounted to the time of infection, was for HIV infection at age 84 years, diagnosed at age 90 years.

**Figure 3:**
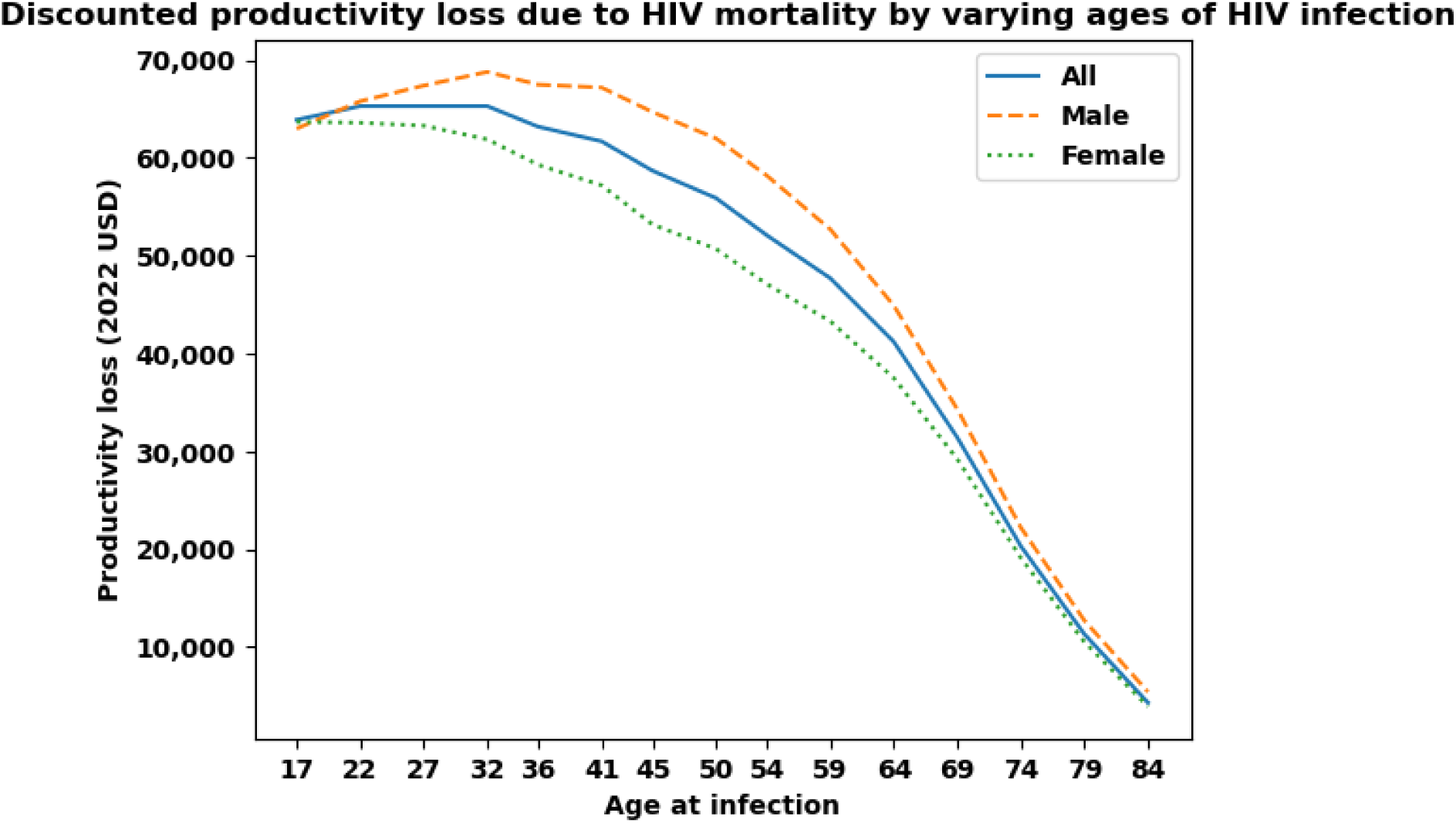
Age-specific productivity loss due to HIV mortality in the United States, discounted to the time of infection and reported in 2022 US dollars.

### Sensitivity Analyses

Varying the percentage of deaths attributable to HIV (*Z*) from 25% to 50% in place of the base case assumption of 30% caused the overall estimated discounted lifetime productivity loss due to HIV mortality per HIV infection to vary from $59,800 to $120,000 (Table 3). Accounting for a small degree of possible heterogeneity in the age at death increased the estimated productivity loss due to HIV mortality per HIV infection for overall by about 54%, from $65,300 to $100,500 (Table 4). This relative impact was similar when stratified by sex.

**Table 3:** Sensitivity analyses of the discounted productivity loss per HIV infection due to HIV mortality (in 2022 US dollars), discounted to the time of infection.

|  | **Percent HIV deaths attributable to HIV (Z)** | **Discounted productivity loss due to HIV mortality per HIV infection ($PL_I$)\*** |  |  |
| --- | --- | --- | --- | --- |
|  |  | **All** | **Male** | **Female** |
| Base case | 30% | \$65,300 | \$67,400 | \$63,300 |
| Varying the percent HIV deaths attributable to HIV |  |  |  |  |
| lower bound | 25% | \$59,800 | \$59,000 | \$60,600 |
| upper bound | 50% | \$120,000 | \$118,400 | \$121,700 |

\*Productivity loss estimates rounded to nearest \$100

**Table 4:** Productivity loss due to HIV mortality, discounted to the time of infection in 2022 US dollars accounting for heterogeneity at life expectancy after HIV diagnosis.

|  | All | Male | Female |
| --- | --- | --- | --- |
| **Base case scenario** |  |  |  |
| Age at death | 66.4 | 66.1 | 66.6 |
| Age at death if not infected | 80.1 | 77.9 | 82.2 |
| Years of life lost | 13.7 | 11.8 | 15.6 |
| Productivity loss due to HIV mortality | \$65,300 | \$67,400 | \$63,300 |
| **Group 1: Those dying 5 years after diagnosis** |  |  |  |
| Age at death | 35 | 35 | 35 |
| Age at death if not infected | 80.1 | 77.9 | 82.2 |
| Years of life lost | 45.1 | 42.9 | 47.2 |
| Productivity loss due to HIV mortality | \$1,013,600 | \$1,139,900 | \$889,500 |
| **Group 2: Those not dying within 5 years of diagnosis** |  |  |  |
| Age at death | 68.0 | 67.7 | 68.2 |
| Age at death if not infected | 80.1 | 77.9 | 82.2 |
| Years of life lost | 12.1 | 10.2 | 14.0 |
| Productivity loss due to HIV mortality | \$53,500 | \$53,500 | \$53,100 |
| **Weighted average of Group 1 and Group 2** |  |  |  |
| Age at death | 66.4 | 66.1 | 66.6 |
| Age at death if not infected | 80.1 | 77.9 | 82.2 |
| Years of life lost | 13.7 | 11.8 | 15.6 |
| Productivity loss due to HIV mortality | \$100,500 | \$106,700 | \$94,100 |
| Percent increase when accounting for<br>heterogeneity | 54% | 58% | 49% |

## Discussion

We estimated, under various scenarios, the productivity loss due to HIV mortality per HIV infection in the United States discounted to the time of HIV infection. In the base case, the overall average discounted lifetime productivity loss due to HIV mortality for an HIV infection diagnosed at age 30 years was estimated to be $65,300 per HIV infection, which was 45% lower than our estimate of $119,700 per HIV infection when applying life expectancy estimates from a 2010 US population-based study restricted to persons with diagnosed HIV. This reduction is primarily due to an approximate 5 years increase in life expectancy among PWH in 2018 from 2010. In addition, our estimate is approximately 91% lower than the most recent estimate ($1,207,222 in 2022 US dollars) of productivity loss of HIV mortality.^14^ This substantial reduction likely reflects improvements in the management and treatment of HIV, especially over the past decade, such as immediate ART initiation^15^ i.e., initiating ART as soon as possible after HIV diagnosis to prevent HIV-associated morbidity and mortality, leading to increased life expectancy after HIV diagnosis.

The base case average productivity loss per HIV infection due to HIV mortality was similar for males ($67,400) and females ($63,300). Although the average number of years of life lost due to HIV mortality is higher for women than men, the annual productivity values are higher for men than for women, and these two differences largely cancel each other out. Results from the sensitivity analysis incorporating heterogeneity in life expectancy shows significantly higher productivity loss from deaths that happen quickly after diagnosis, which can be because of a late diagnosis of an HIV infection in an advanced stage or absence of HIV treatment.

Our findings further highlight the impact of advancements in ART and importantly more PWH on ART on productivity loss as the estimated productivity loss per HIV infection due to HIV mortality was notably lower, both overall and stratified by sex, when we assumed that all people diagnosed with HIV would be treated with ART. Our estimated overall productivity loss due to HIV mortality after a diagnosis at age 30 years, applying life expectancy data from cohorts on ART initiating and maintaining ART between 2015-2019 was 83% lower compared to the base case estimate.

Our base case estimated productivity loss per HIV infection due to HIV mortality of $65,300 is about 14% of the estimated lifetime direct medical cost per HIV infection ($481,100 in 2022 USD).^10^ Still, the productivity losses are substantial and represent an important component of the economic burden of HIV. A productivity loss of $65,300 per HIV infection, as reported in our base case, when multiplied by a recent estimate of 31,800 incident HIV infections in 2022 in the United States^8,42^, suggests that incident HIV infections in 2022 imposed a productivity loss due to HIV mortality of about $2.1 billion. Estimates stratified by sex and by age can also be applied to corresponding incidence estimates to calculate the productivity loss from specific groups. For example, in 2022, the projected productivity loss of HIV mortality of new HIV infections in the age group 35-44 years (6,700) is within the range of $413-$423 million using the corresponding age specific productivity loss estimates.

We addressed a gap in the literature by providing updated estimates of HIV mortality-related productivity losses. These estimates can be useful in combination with the recent lifetime treatment cost per HIV infection^10^ to estimate the aggregated economic burden of a new HIV infection and apply that to incidence infections to assess the total economic burden of new HIV infections that happen each year. These estimates should help policy makers to understand the total burden of new HIV infections and plan for HIV prevention and care strategies accordingly.

### Limitations

There are several limitations to this study. First, our findings are sensitive to the key inputs (e.g., percentage of deaths among PWH attributable to HIV, life expectancy) in our analysis, and these key inputs are subject to considerable variability. To address this limitation, we conducted sensitivity analyses to illustrate how the results change when varying these key assumptions.

However, we did not vary other input parameters (e.g., years from HIV infection to diagnosis) due to low reported variability that minimally impacts the lifetime productivity lost estimates. Second, in the calculation of years of life lost estimates for PWH on ART from a collaborative cohort study in Europe and North America compared to general population life expectancy, we used the life expectancy data of the US general population. This approach may have over– or under-estimated the years of life lost if the life expectancy of general population of the other countries included in the cohort studies is lower or higher compared to that of the US general population. Third, for simplicity and data availability, the scope of our study was limited to discounted productivity loss due to HIV mortality per HIV infection for overall and stratified by sex only. We did not attempt to estimate the mortality-related productivity loss of HIV by transmission groups, by race/ethnicity or by disease stage at HIV diagnosis as the key input parameter, annual productivity data, is not available by these stratifications. Fourth, in the absence of life expectancy data stratified by HIV-attributable and non-HIV-attributable deaths among PWH, we employed average life years lost estimates for all PWH in our productivity loss estimates and adjusted that by the percentages of deaths attributable to HIV. Fifth, we made the simplifying assumption that productivity in people with HIV is the same as for people without HIV. However, an extensive literature documents differences in many factors affecting productivity, such as labor force participation rates, physical and mental health-related quality of life. ^43,44^ Hence, those who prefer to assume differential productivity by HIV status can adjust our results proportionally, or replicate our approach using differential annual productivity, in accordance with their assumptions about productivity among PWH. Sixth, our study was limited to productivity loss due to HIV mortality and did not include productivity loss of HIV morbidity, such as time missed from work due to HIV-attributable illness or disability.

## Conclusion

In conclusion, our study provides a useful update of the estimated lifetime productivity loss per HIV infection associated with HIV mortality. We estimated a 45% reduction in the lifetime discounted productivity loss per HIV infection due to HIV mortality over the past decade. We also observed reductions in lifetime productivity loss due to HIV mortality with advancements in treatment, highlighting the positive impact of improved HIV care in the United States. Findings from this study can help in quantifying the total economic burden of HIV in the United States and can be used in cost-effectiveness analyses of HIV prevention interventions. Future analyses could expand on this work to include productivity losses associated with HIV morbidity and to examine productivity losses among additional subpopulations.

## Data Availability

All data produced in the present work are contained in the manuscript

## Supplementary Materials

**Table S1:** Age-specific annual mean productivity of United States residents by sex and productivity type in 2022 US dollars.

| Age | Male |  |  | Female |  |  | All |  |  |
| --- | --- | --- | --- | --- | --- | --- | --- | --- | --- |
|  | Market | Non-market | Total | Market | Non-market | Total | Market | Non-market | Total |
| 17 | \$2,573 | \$7,623 | \$10,196 | \$2,546 | \$11,243 | \$13,790 | \$2,559 | \$9,387 | \$11,947 |
| 18 | \$5,307 | \$7,297 | \$12,603 | \$4,723 | \$10,512 | \$15,233 | \$5,023 | \$8,875 | \$13,897 |
| 19 | \$9,494 | \$7,588 | \$17,082 | \$7,748 | \$11,697 | \$19,445 | \$8,643 | \$9,587 | \$18,229 |
| 20 | \$13,724 | \$9,212 | \$22,935 | \$10,695 | \$15,047 | \$25,742 | \$12,255 | \$12,061 | \$24,315 |
| 21 | \$17,908 | \$10,539 | \$28,448 | \$14,051 | \$17,401 | \$31,452 | \$16,048 | \$13,845 | \$29,891 |
| 22 | \$22,167 | \$12,048 | \$34,215 | \$17,784 | \$19,974 | \$37,760 | \$20,049 | \$15,836 | \$35,885 |
| 23 | \$27,064 | \$13,703 | \$40,767 | \$22,158 | \$22,464 | \$44,622 | \$24,675 | \$17,993 | \$42,668 |
| 24 | \$33,554 | \$15,160 | \$48,713 | \$27,330 | \$24,944 | \$52,274 | \$30,497 | \$19,969 | \$50,466 |
| 25 | \$39,566 | \$16,209 | \$55,775 | \$31,816 | \$27,671 | \$59,486 | \$35,753 | \$21,846 | \$57,599 |
| 26 | \$44,948 | \$16,903 | \$61,851 | \$35,525 | \$30,507 | \$66,032 | \$40,301 | \$23,602 | \$63,903 |
| 27 | \$48,782 | \$17,511 | \$66,293 | \$37,706 | \$33,315 | \$71,022 | \$43,298 | \$25,333 | \$68,632 |
| 28 | \$52,428 | \$18,212 | \$70,640 | \$39,524 | \$35,892 | \$75,416 | \$46,026 | \$27,003 | \$73,029 |
| 29 | \$56,333 | \$18,984 | \$75,317 | \$40,996 | \$38,326 | \$79,321 | \$48,731 | \$28,578 | \$77,310 |
| 30 | \$60,260 | \$20,388 | \$80,647 | \$42,700 | \$40,605 | \$83,304 | \$51,545 | \$30,378 | \$81,923 |
| 31 | \$64,165 | \$22,469 | \$86,634 | \$44,164 | \$42,580 | \$86,745 | \$54,244 | \$32,492 | \$86,737 |
| 32 | \$67,748 | \$25,174 | \$92,921 | \$45,481 | \$44,042 | \$89,522 | \$56,661 | \$34,523 | \$91,184 |
| 33 | \$70,408 | \$27,966 | \$98,374 | \$46,421 | \$44,815 | \$91,236 | \$58,451 | \$36,386 | \$94,838 |
| 34 | \$73,751 | \$29,689 | \$103,440 | \$47,776 | \$45,033 | \$92,809 | \$60,791 | \$37,359 | \$98,150 |
| 35 | \$76,589 | \$30,747 | \$107,337 | \$49,166 | \$45,107 | \$94,271 | \$62,879 | \$37,889 | \$100,768 |
| 36 | \$80,370 | \$30,728 | \$111,098 | \$50,129 | \$44,980 | \$95,109 | \$65,257 | \$37,890 | \$103,147 |
| 37 | \$82,793 | \$30,679 | \$113,472 | \$50,600 | \$45,074 | \$95,672 | \$66,586 | \$37,867 | \$104,453 |
| 38 | \$85,700 | \$30,566 | \$116,266 | \$51,480 | \$44,816 | \$96,297 | \$68,517 | \$37,807 | \$106,325 |
| 39 | \$87,342 | \$30,530 | \$117,871 | \$52,256 | \$44,270 | \$96,526 | \$69,651 | \$37,383 | \$107,035 |
| 40 | \$89,368 | \$30,551 | \$119,920 | \$53,131 | \$43,157 | \$96,288 | \$71,219 | \$36,926 | \$108,145 |
| 41 | \$90,383 | \$30,786 | \$121,169 | \$53,321 | \$41,714 | \$95,036 | \$71,759 | \$36,230 | \$107,989 |
| 42 | \$92,193 | \$30,713 | \$122,907 | \$53,721 | \$39,890 | \$93,613 | \$72,825 | \$35,337 | \$108,163 |
| 43 | \$94,035 | \$30,037 | \$124,072 | \$54,229 | \$38,342 | \$92,570 | \$73,881 | \$34,255 | \$108,137 |
| 44 | \$95,895 | \$28,604 | \$124,499 | \$54,880 | \$36,891 | \$91,770 | \$75,161 | \$32,803 | \$107,965 |
| 45 | \$97,105 | \$26,711 | \$123,816 | \$55,724 | \$35,889 | \$91,612 | \$76,219 | \$31,317 | \$107,535 |
| 46 | \$97,272 | \$25,214 | \$122,486 | \$55,594 | \$34,662 | \$90,256 | \$76,261 | \$29,990 | \$106,251 |
| 47 | \$97,545 | \$24,380 | \$121,925 | \$55,450 | \$33,442 | \$88,893 | \$76,294 | \$28,956 | \$105,250 |
| 48 | \$97,540 | \$24,136 | \$121,676 | \$55,063 | \$32,129 | \$87,192 | \$76,047 | \$28,165 | \$104,211 |
| 49 | \$95,643 | \$23,726 | \$119,369 | \$54,815 | \$30,891 | \$85,708 | \$74,950 | \$27,372 | \$102,324 |
| 50 | \$94,241 | \$23,289 | \$117,529 | \$54,481 | \$29,740 | \$84,221 | \$74,053 | \$26,564 | \$100,616 |
| 51 | \$93,494 | \$22,583 | \$116,077 | \$53,759 | \$28,646 | \$82,406 | \$73,308 | \$25,655 | \$98,963 |
| 52 | \$94,394 | \$22,034 | \$116,428 | \$53,695 | \$27,815 | \$81,509 | \$73,732 | \$24,980 | \$98,712 |
| 53 | \$93,964 | \$21,156 | \$115,120 | \$53,338 | \$27,331 | \$80,669 | \$73,288 | \$24,284 | \$97,572 |
| 54 | \$91,546 | \$19,937 | \$111,482 | \$52,645 | \$27,170 | \$79,814 | \$71,737 | \$23,630 | \$95,367 |
| 55 | \$89,606 | \$18,801 | \$108,407 | \$51,100 | \$27,156 | \$78,256 | \$69,980 | \$23,068 | \$93,048 |
| 56 | \$87,839 | \$17,771 | \$105,610 | \$49,654 | \$27,303 | \$76,955 | \$68,306 | \$22,612 | \$90,918 |
| 57 | \$86,728 | \$17,178 | \$103,907 | \$48,485 | \$27,637 | \$76,122 | \$67,115 | \$22,576 | \$89,691 |
| 58 | \$84,588 | \$17,082 | \$101,671 | \$47,083 | \$28,026 | \$75,109 | \$65,236 | \$22,715 | \$87,952 |
| 59 | \$81,711 | \$17,277 | \$98,988 | \$44,859 | \$28,622 | \$73,481 | \$62,667 | \$23,142 | \$85,810 |
| 60 | \$78,371 | \$17,745 | \$96,116 | \$41,922 | \$29,113 | \$71,035 | \$59,434 | \$23,641 | \$83,074 |
| 61 | \$74,053 | \$18,321 | \$92,374 | \$38,905 | \$29,366 | \$68,271 | \$55,678 | \$24,098 | \$79,777 |
| 62 | \$68,746 | \$19,126 | \$87,872 | \$35,410 | \$29,684 | \$65,094 | \$51,272 | \$24,690 | \$75,961 |
| 63 | \$62,876 | \$19,979 | \$82,854 | \$31,711 | \$29,739 | \$61,448 | \$46,521 | \$25,077 | \$71,598 |
| 64 | \$55,931 | \$20,836 | \$76,767 | \$27,384 | \$29,601 | \$56,986 | \$40,942 | \$25,436 | \$66,378 |
| 65 | \$49,489 | \$21,758 | \$71,246 | \$23,414 | \$29,420 | \$52,832 | \$35,787 | \$25,808 | \$61,595 |
| 66 | \$42,742 | \$22,166 | \$64,908 | \$19,549 | \$29,302 | \$48,852 | \$30,534 | \$25,898 | \$56,433 |
| 67 | \$36,978 | \$22,213 | \$59,191 | \$16,325 | \$29,228 | \$45,553 | \$26,078 | \$25,913 | \$51,991 |
| 68 | \$32,246 | \$22,134 | \$54,380 | \$13,615 | \$29,355 | \$42,970 | \$22,370 | \$25,993 | \$48,363 |
| 69 | \$27,672 | \$22,134 | \$49,806 | \$11,453 | \$29,396 | \$40,849 | \$19,053 | \$25,975 | \$45,027 |
| 70 | \$24,724 | \$22,105 | \$46,830 | \$9,438 | \$29,290 | \$38,727 | \$16,565 | \$25,919 | \$42,483 |
| 71 | \$21,891 | \$22,011 | \$43,902 | \$8,031 | \$29,011 | \$37,042 | \$14,431 | \$25,805 | \$40,236 |
| 72 | \$19,687 | \$22,101 | \$41,787 | \$6,827 | \$28,542 | \$35,369 | \$12,730 | \$25,591 | \$38,321 |
| 73 | \$17,531 | \$22,100 | \$39,630 | \$6,057 | \$28,010 | \$34,065 | \$11,321 | \$25,281 | \$36,602 |
| 74 | \$15,388 | \$22,067 | \$37,455 | \$5,205 | \$27,308 | \$32,512 | \$9,857 | \$24,914 | \$34,771 |
| 75 | \$13,441 | \$21,898 | \$35,339 | \$4,548 | \$26,624 | \$31,172 | \$8,572 | \$24,491 | \$33,063 |
| 76 | \$11,760 | \$21,523 | \$33,282 | \$4,098 | \$25,693 | \$29,793 | \$7,536 | \$23,828 | \$31,363 |
| 77 | \$10,369 | \$21,107 | \$31,478 | \$3,470 | \$24,781 | \$28,251 | \$6,530 | \$23,142 | \$29,671 |
| 78 | \$9,877 | \$20,761 | \$30,638 | \$3,036 | \$23,820 | \$26,855 | \$6,049 | \$22,486 | \$28,535 |
| 79 | \$8,707 | \$20,073 | \$28,781 | \$2,459 | \$22,506 | \$24,965 | \$5,169 | \$21,438 | \$26,606 |
| 80 | \$7,533 | \$19,232 | \$26,765 | \$2,075 | \$21,232 | \$23,307 | \$4,429 | \$20,385 | \$24,814 |
| 81 | \$6,208 | \$18,387 | \$24,594 | \$1,679 | \$19,865 | \$21,545 | \$3,597 | \$19,232 | \$22,828 |
| 82 | \$5,388 | \$17,311 | \$22,700 | \$1,407 | \$18,546 | \$19,954 | \$3,096 | \$18,031 | \$21,127 |
| 83 | \$4,632 | \$16,214 | \$20,847 | \$1,283 | \$17,104 | \$18,387 | \$2,669 | \$16,725 | \$19,395 |
| **84** | \$4,237 | \$14,826 | \$19,063 | \$1,208 | \$15,393 | \$16,602 | \$2,445 | \$15,171 | \$17,616 |
| **85** | \$3,739 | \$13,581 | \$17,320 | \$1,089 | \$13,648 | \$14,737 | \$2,124 | \$13,621 | \$15,744 |
| **86** | \$3,817 | \$12,192 | \$16,009 | \$882 | \$12,287 | \$13,169 | \$2,019 | \$12,251 | \$14,270 |
| **87** | \$3,517 | \$10,763 | \$14,281 | \$754 | \$10,817 | \$11,571 | \$1,788 | \$10,796 | \$12,584 |
| **88** | \$3,482 | \$9,552 | \$13,035 | \$701 | \$9,285 | \$9,988 | \$1,721 | \$9,382 | \$11,101 |
| **89** | \$2,887 | \$8,184 | \$11,073 | \$571 | \$7,911 | \$8,482 | \$1,372 | \$8,008 | \$9,379 |
| **90** | \$2,223 | \$6,778 | \$9,001 | \$422 | \$6,428 | \$6,850 | \$1,025 | \$6,541 | \$7,566 |
| **91** | \$2,514 | \$5,497 | \$8,011 | \$348 | \$5,176 | \$5,524 | \$1,022 | \$5,279 | \$6,300 |
| **92** | \$2,105 | \$4,584 | \$6,688 | \$321 | \$4,153 | \$4,475 | \$866 | \$4,285 | \$5,151 |
| **93** | \$2,367 | \$3,562 | \$5,929 | \$352 | \$3,251 | \$3,604 | \$956 | \$3,341 | \$4,297 |
| **94** | \$1,585 | \$2,561 | \$4,146 | \$335 | \$2,329 | \$2,666 | \$700 | \$2,397 | \$3,097 |
| **95** | \$1,690 | \$1,816 | \$3,506 | \$327 | \$1,646 | \$1,973 | \$724 | \$1,696 | \$2,420 |
| **96** | \$1,352 | \$1,191 | \$2,544 | \$261 | \$1,078 | \$1,339 | \$579 | \$1,111 | \$1,690 |
| **97** | \$1,015 | \$696 | \$1,711 | \$196 | \$629 | \$826 | \$4 | \$649 | \$1,083 |
| **98** | \$676 | \$338 | \$1,015 | \$130 | \$305 | \$435 | \$290 | \$315 | \$605 |
| **99** | \$507 | \$182 | \$689 | \$98 | \$163 | \$262 | \$217 | \$169 | \$387 |

Source: Personal communication with the corresponding author of Grosse SD, Krueger KV, Pike J.
Estimated annual and lifetime labor productivity in the United States, 2016: implications for economic evaluations. J Med Econ. 2019 Jun;22(6):501-508.

**Table S2:** Age at HIV infection, median time from HIV infection to diagnosis, age at HIV diagnosis, remaining life expectancy at HIV diagnosis, and number of life years lost per person with HIV.

| Age at HIV infection | Median time from HIV infection to diagnosis (M) | Age at HIV diagnosis | Remaining Life expectancy (in years) at HIV diagnosis (*E*) |  |  | Number of years of life lost among PWH (*L*) |  |  |
| --- | --- | --- | --- | --- | --- | --- | --- | --- |
|  |  |  | All | Male | Female | All | Male | Female |
| 17 | 3 | 20 | 44.8 | 44.9 | 44.6 | 14.8 | 12.2 | 17.3 |
| 22 | 3 | 25 | 40.5 | 40.4 | 40.6 | 14.3 | 12.0 | 16.5 |
| 27 | 3 | 30 | 36.4 | 36.1 | 36.6 | 13.7 | 11.8 | 15.6 |
| 32 | 3 | 35 | 32.3 | 31.8 | 32.8 | 13.1 | 11.5 | 14.7 |
| 36 | 4 | 40 | 28.4 | 27.7 | 29.0 | 12.5 | 11.1 | 13.8 |
| 41 | 4 | 45 | 24.5 | 23.6 | 25.3 | 11.7 | 10.6 | 12.8 |
| 45 | 5 | 50 | 20.7 | 19.7 | 21.7 | 11.0 | 10.1 | 11.8 |
| 50 | 5 | 55 | 17.1 | 16.1 | 18.1 | 10.3 | 9.6 | 11.0 |
| 54 | 6 | 60 | 13.6 | 12.6 | 14.5 | 9.8 | 9.2 | 10.3 |
| 59 | 6 | 65 | 10.3 | 9.5 | 11.1 | 9.1 | 8.6 | 9.6 |
| 64 | 6 | 70 | 7.4 | 6.8 | 8.0 | 8.3 | 7.8 | 8.7 |
| 69 | 6 | 75 | 5.1 | 4.6 | 5.5 | 7.2 | 6.7 | 7.6 |
| 74 | 6 | 80 | 3.4 | 3.1 | 3.6 | 5.8 | 5.4 | 6.2 |
| 79 | 6 | 85 | 1.9 | 1.8 | 2.0 | 4.6 | 4.2 | 5.0 |
| 84 | 6 | 90 | 1.1 | 1.0 | 1.1 | 3.4 | 3.1 | 3.7 |

Source: Siddiqi AE, Johnson AS, Hernandez A, Hu X, Song R. Life Expectancy after HIV Diagnosis 2008-
2018, United States [Abstract 761]. presented at: Conference on Retroviruses and Opportunistic
Infections; February 12-16, 2022; Virtual.

**Table S3:** Productivity loss due to HIV mortality in the United States in 2022 US dollars, discounted to the time of infection by varying age of HIV infection.

| Age at<br>HIV<br>infection | Productivity loss per HIV infection due to HIV mortality ($PL_I$)* | | | | | | | | |
| --- | --- | --- | --- | --- | --- | --- | --- | --- | --- |
|  | All |  |  | Male |  |  | Female |  |  |
|  | Market | Non-Market | Total | Market | Non-Market | Total | Market | Non-Market | Total |
| 17 | 25,800 | 38,100 | 63,900 | 34,800 | 28,300 | 63,000 | 16,500 | 47,300 | 63,700 |
| 22 | 25,100 | 40,200 | 65,300 | 35,400 | 30,400 | 65,800 | 14,900 | 48,700 | 63,600 |
| 27 | 23,600 | 41,700 | 65,300 | 34,800 | 32,600 | 67,400 | 13,500 | 49,700 | 63,300 |
| 32 | 22,200 | 43,100 | 65,300 | 34,200 | 34,600 | 68,800 | 11,800 | 50,100 | 61,900 |
| 36 | 19,900 | 43,300 | 63,200 | 32,000 | 35,500 | 67,500 | 10,100 | 49,300 | 59,300 |
| 41 | 18,100 | 43,600 | 61,700 | 30,400 | 36,800 | 67,200 | 8,700 | 48,500 | 57,200 |
| 45 | 15,900 | 42,800 | 58,700 | 27,600 | 37,000 | 64,700 | 7,300 | 46,000 | 53,200 |
| 50 | 13,800 | 42,100 | 55,900 | 24,500 | 37,500 | 62,000 | 6,200 | 44,500 | 50,700 |
| 54 | 11,600 | 40,600 | 52,100 | 21,000 | 37,100 | 58,200 | 5,200 | 41,900 | 47,100 |
| 59 | 9,500 | 38,200 | 47,700 | 17,000 | 35,700 | 52,700 | 4,300 | 39,100 | 43,300 |
| 64 | 7,200 | 34,000 | 41,200 | 12,800 | 32,100 | 44,900 | 3,200 | 34,300 | 37,500 |
| 69 | 4,700 | 26,700 | 31,400 | 8,500 | 25,800 | 34,300 | 2,100 | 27,000 | 29,200 |
| 74 | 2,900 | 17,500 | 20,300 | 5,100 | 17,100 | 22,200 | 1,300 | 17,800 | 19,100 |
| 79 | 1,700 | 9,600 | 11,300 | 3,200 | 9,500 | 12,700 | 700 | 9,800 | 10,500 |
| 84 | 800 | 3,500 | 4,300 | 1,800 | 3,600 | 5,400 | 300 | 3,600 | 3,900 |

